# Knowledge, Attitudes, and Barriers to Oncofertility Services Among Cancer Patients Requiring Chemoradiotherapy: A Systematic Review

**DOI:** 10.1101/2025.08.13.25333645

**Authors:** James Njiru, Poli A. Philippe, Richard Mogeni, Elly Odongo, Omenge O Elkanah

**Author notes:** **Corresponding author**: Dr. Poli Philippe Amubuomombe, MD, Directorate of Reproductive Health, Moi Teaching and Referral Hospital, P.O Box 3-30100 Eldoret, Kenya.

## Abstract

**Background:** Cancer treatments such as chemotherapy and radiotherapy can impair fertility, making fertility preservation critical for reproductive-aged patients. Despite clinical guidelines supporting fertility counseling, uptake of oncofertility services remains low.

**Objective:** This systematic review aims to examine the current evidence on cancer patients’ knowledge, attitudes, and perceived barriers toward oncofertility services.

**Methods:** A systematic search of PubMed, Hinari, EBSCOhost, and Google Scholar was conducted for English-language studies published between January 2014 and August 2024. Eligible studies included quantitative, qualitative, or mixed-method designs focusing on fertility preservation knowledge, attitudes, or barriers among patients undergoing cancer treatment. Data were synthesized narratively.

**Results:** Thirteen studies met inclusion criteria. Most patients had limited knowledge of fertility preservation, often due to insufficient counselling. Despite this, many expressed strong interest in fertility preservation. Barriers included lack of information, high costs, systemic inefficiencies, cultural beliefs, and psychological distress.

**Conclusion:** Integration of fertility counselling into oncology care, greater provider training, financial support policies, and culturally sensitive interventions are essential to improve access to oncofertility services.

## Introduction

The increasing incidence of cancer among individuals of reproductive age has brought renewed attention to survivorship issues, particularly the preservation of fertility [1, 2]. With improvements in early detection and therapeutic advancements, survival rates have significantly improved for many cancers, including breast, lymphoma, and gynecologic malignancies [3, 4, 5, 6]. However, these treatments, while lifesaving, often compromise reproductive potential through gonadotoxic effects. Chemotherapy can damage germ cells and suppress gonadal function in both sexes [7, 8, 9]. Similarly, radiotherapy, especially when directed at the pelvis or abdomen, can result in irreversible impairment of ovarian or testicular function [10, 11, 12]. Hormonal changes and structural damage following treatment can further reduce the likelihood of future conception [13, 14].

Beyond the physiological impacts, infertility as a result of cancer therapy has profound psychosocial implications. Numerous studies have documented the psychological distress that accompanies loss of fertility, including depression, anxiety, diminished self-esteem, strained intimate relationships, and overall reduced quality of life [15, 16, 17]. For many young adults and adolescents, the ability to conceive is not only a matter of biological function but also a key aspect of personal identity, life aspirations, and cultural expectations [16, 18]. Fertility considerations may influence patients’ choices regarding treatment regimens, willingness to delay therapy, or engage in complex multidisciplinary consultations [15].

To address these challenges, the field of oncofertility has emerged at the intersection of oncology and reproductive medicine. It seeks to safeguard future fertility through a range of interventions, including sperm cryopreservation, oocyte and embryo freezing, ovarian tissue preservation, testicular tissue cryopreservation, and the use of gonadotropin-releasing hormone (GnRH) analogues to protect ovarian function during treatment [1, 20]. These technologies have progressed rapidly over the past two decades and are increasingly supported by national and international clinical guidelines [1, 2, 21]. Organizations such as the American Society of Clinical Oncology (ASCO) and the European Society for Medical Oncology (ESMO) recommend that fertility preservation be discussed as early as possible after diagnosis and before initiation of treatment [1, 2].

Despite these advancements, global uptake of fertility preservation services remains suboptimal [1, 22, 23]. Many patients are not adequately informed about fertility risks, nor are they referred to specialists in time to take advantage of available interventions [15]. Several studies have reported that oncology teams often omit fertility discussions, citing time constraints, lack of knowledge, absence of clear referral pathways, or assumptions about patient priorities. In many cases, patients report having to advocate for themselves or learning about fertility options only after therapy has commenced—when opportunities for preservation may have already been lost [22, 24].

Barriers to fertility preservation are multifactorial. These include informational deficits, financial constraints, structural limitations within healthcare systems, cultural and religious beliefs, and the psychological readiness of the patient [15, 22, 25]. Understanding how these factors intersect is essential for designing targeted interventions that improve access and equity in oncofertility care.

This systematic review synthesizes current evidence on cancer patients’ knowledge of fertility preservation, their attitudes toward such services, and the barriers they face in accessing care. By exploring perspectives from diverse geographical and cultural contexts, this review aims to provide actionable insights to inform clinical guidelines, policy reforms, and patient-centered oncofertility programs.

## Methods

### Study Design and Protocol Registration

This review employed a systematic review methodology and was conducted in accordance with the Preferred Reporting Items for Systematic Reviews and Meta-Analyses (PRISMA) guidelines 2020 [26]. Although not registered prospectively, the review protocol followed established best practices to ensure transparency, reproducibility, and rigor. The approach was suitable for integrating quantitative and qualitative evidence related to patient perspectives on fertility preservation.

### Databases and Search Strategy

A structured literature search was performed in August 2024 using four major databases: PubMed, Hinari, EBSCOhost, and Google Scholar, as summarized in Figure 1. The search strategy was developed in collaboration with a medical librarian and included combinations of keywords and MeSH terms: “oncofertility,” “fertility preservation,” “cancer,” “chemotherapy,” “radiotherapy,” “patient knowledge,” “attitudes,” “barriers,” and “access.” Boolean operators (AND/OR) were used to refine the search. Additional hand-searching of references from included articles ensured comprehensive coverage.

**Figure 1.**
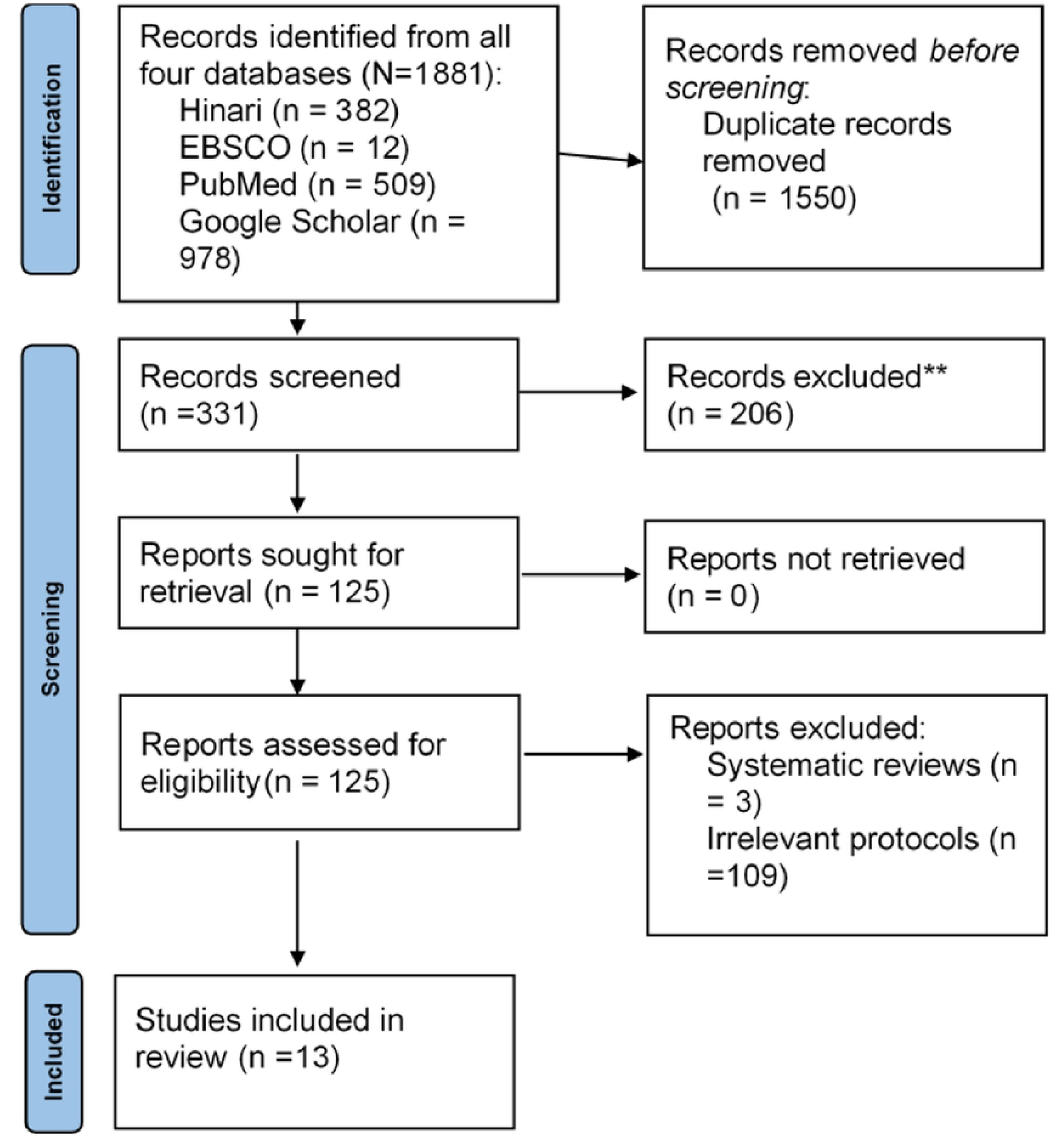

### Inclusion Criteria

Studies were eligible for inclusion if they: (1) involved cancer patients aged 18–45 undergoing or scheduled to undergo chemotherapy and/or radiotherapy; (2) assessed any aspect of knowledge, attitudes, or barriers related to fertility preservation; (3) were peer-reviewed and published in English between January 2014 and August 2024; and (4) used qualitative, quantitative, or mixed-method designs.

### Exclusion Criteria

Excluded studies focused exclusively on healthcare providers, pediatric patients, or lacked sufficient methodological detail for quality appraisal.

### Study Screening and Data Extraction

Two independent reviewers screened titles and abstracts using Rayyan QCRI and retrieved full texts for potentially eligible articles. A data extraction form was designed to collect key information: author, year, country, study design, sample size and demographics, type of cancer, type of fertility preservation discussed, outcomes related to knowledge, attitudes, and barriers, and key conclusions. Discrepancies were resolved by consensus or through a third reviewer.

### Quality Appraisal

The methodological quality of each study was assessed using the Mixed Methods Appraisal Tool (MMAT), 2018 version. Each study was rated based on design-specific criteria (qualitative, quantitative, or mixed methods). Scores were reported as percentages, and only studies achieving a score of 75% or above were included in the final synthesis to maintain methodological robustness. Table 1 summarizes the MMAT quality appraisal exercise results.

**Table 1:** MMAT quality appraisal exercise results.

| Qualitative studies | MMAT scores (%) | 1.1 Are the sources of qualitative data (archives, | 1.2 Is the process for analysing qualitative | 1.3 Is appropriate consideration give to how | 1.4. Is appropriate consideration given to how |
| --- | --- | --- | --- | --- | --- |

|  |  | **documents, informants, observations) relevant to address the research question (objective)?** | **data relevant to address the research question objective?** | **findings relate to the context e.g. the setting, in which the data were collected?** | **findings relate to researchers' influence, e.g. through their interactions with participants?** |
| --- | --- | --- | --- | --- | --- |
| Salsman (2021) | 75 | X | X | X |  |
| Lara del Valle (2022) | 100 | X | X | X | X |
| Niemasik (2012) | 75 | X | X | X |  |
| Taniskidou (2024) | 75 | X | X | X |  |

| Quantitative studies descriptive | (%) | 4.1. Is the sampling strategy relevant to address the quantitative research question (quantitative aspect of the mixed methods question)? | 4.2. Is the sample representative of the population understudy? | 4.3. Are measurements appropriate (clear origin, or validity known, or standard instrument)? | 4.4. Is there an acceptable response rate (60% or above)? |
| --- | --- | --- | --- | --- | --- |
| Stiner (2019) | 75 | X |  | X | X |
| Cherven (2016) | 75 | X | X | X |  |
| Urech (2018) | 75 | X |  | X | X |
| Buyuk (2019) | 75 | X |  | X | X |
| Ko (2023) | 100 | X | X | X | X |
| Zhang (2019) | 100 | X | X | X | X |
| Abusanad (2022) | 100 | X | X | X | X |
| Edge (2005) | 75 | X |  | X | X |
| Zhang (2019) | 100 | X | X | X | X |

### Synthesis Approach

Due to heterogeneity in study design and outcomes, a meta-analysis was not conducted. Instead, a narrative synthesis approach was adopted. Findings were organized under three thematic domains: (1) knowledge of fertility preservation, (2) attitudes and preferences, and (3) barriers to access. Subthemes and illustrative examples were integrated to enhance interpretability and contextual understanding.

## Results

A total of 13 studies were included in this systematic review, spanning countries such as the United States, Canada, Iran, Australia, and the United Kingdom. The studies employed various methodologies including qualitative interviews (n=5), quantitative surveys (n=6), and mixed-methods approaches (n=2). Across all studies, common themes emerged under three main domains: knowledge of fertility preservation, attitudes and preferences, and barriers to access. Table 2 summarizes studies included in this systematic review.

**Table 2:**
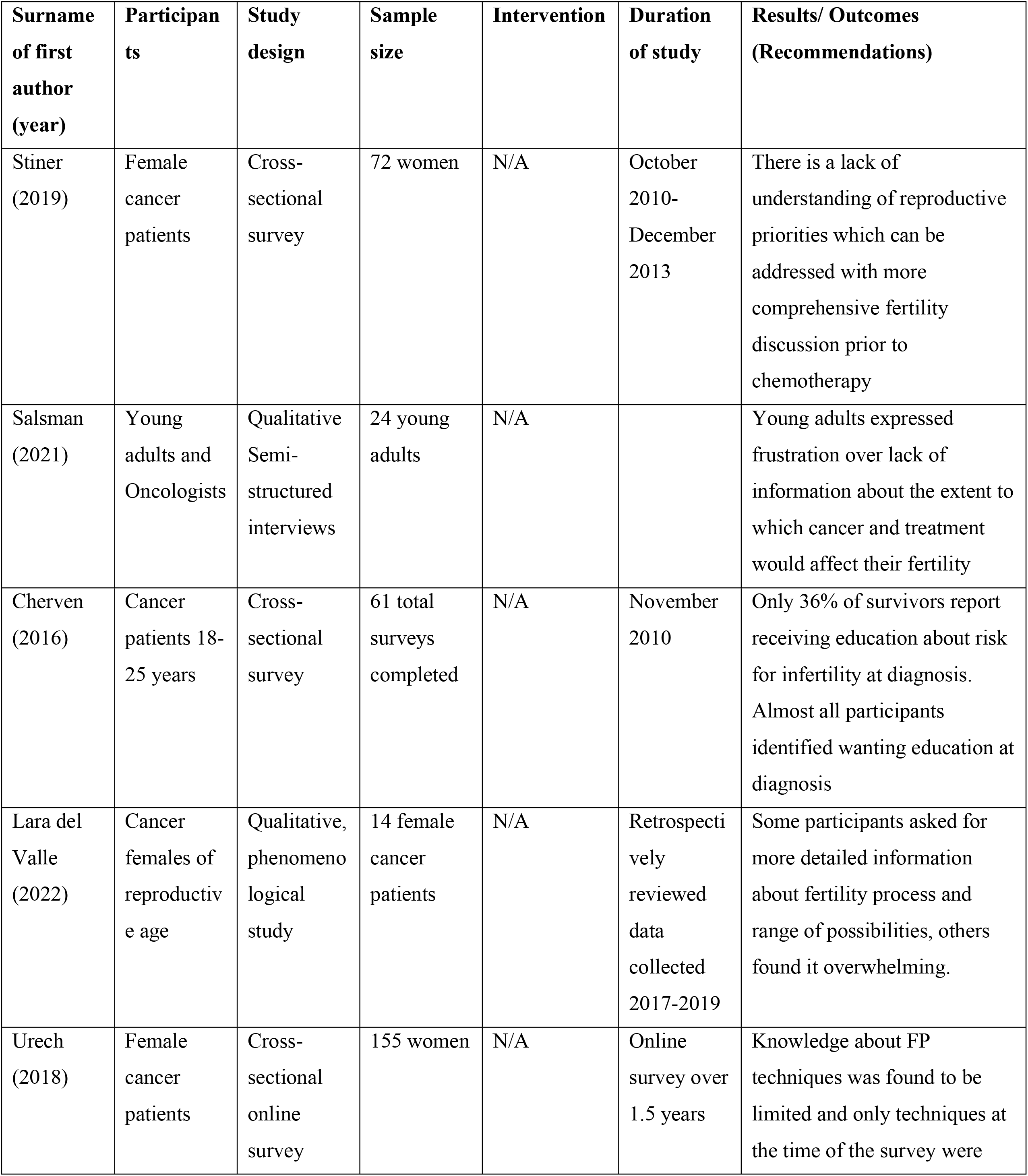
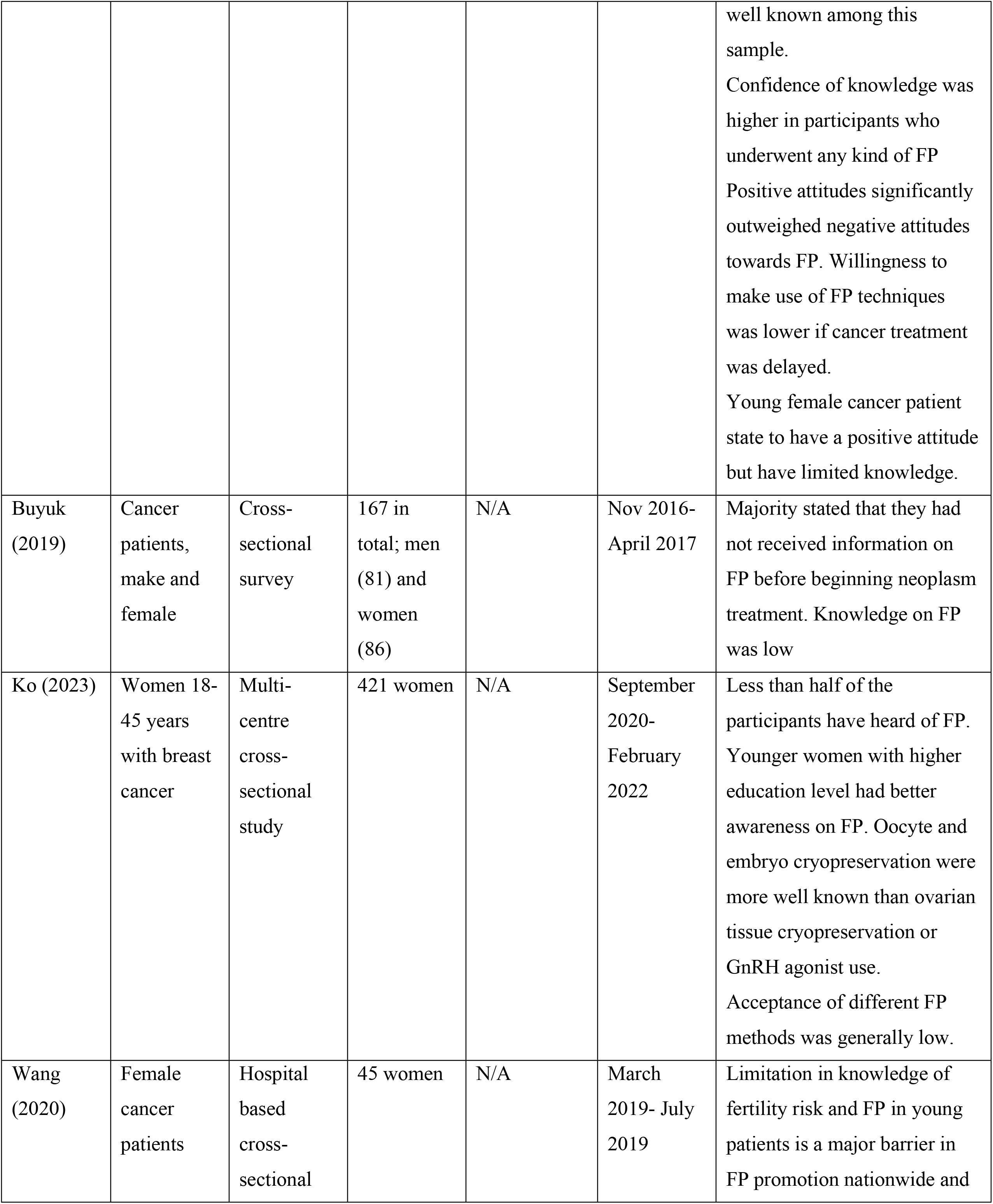
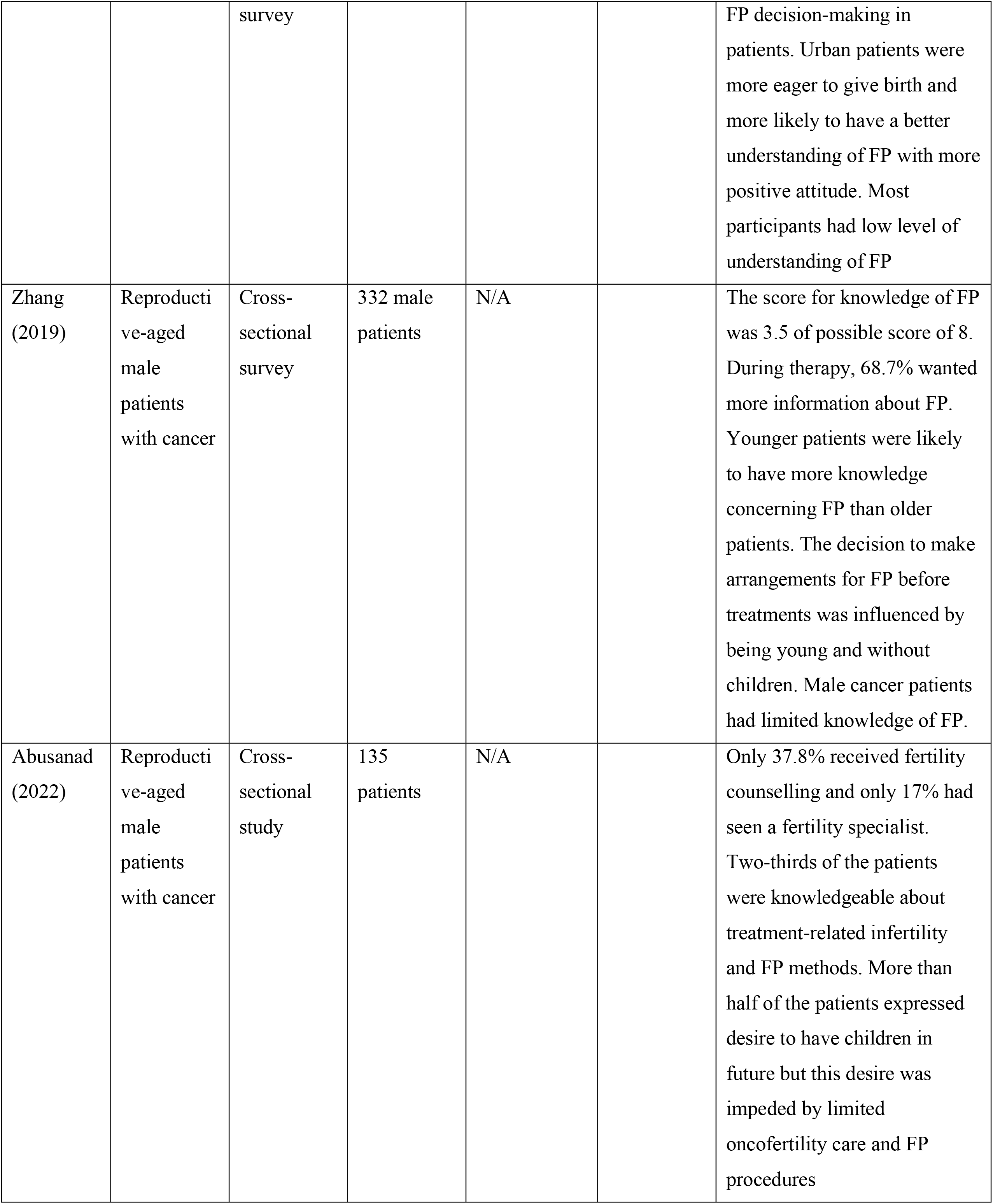
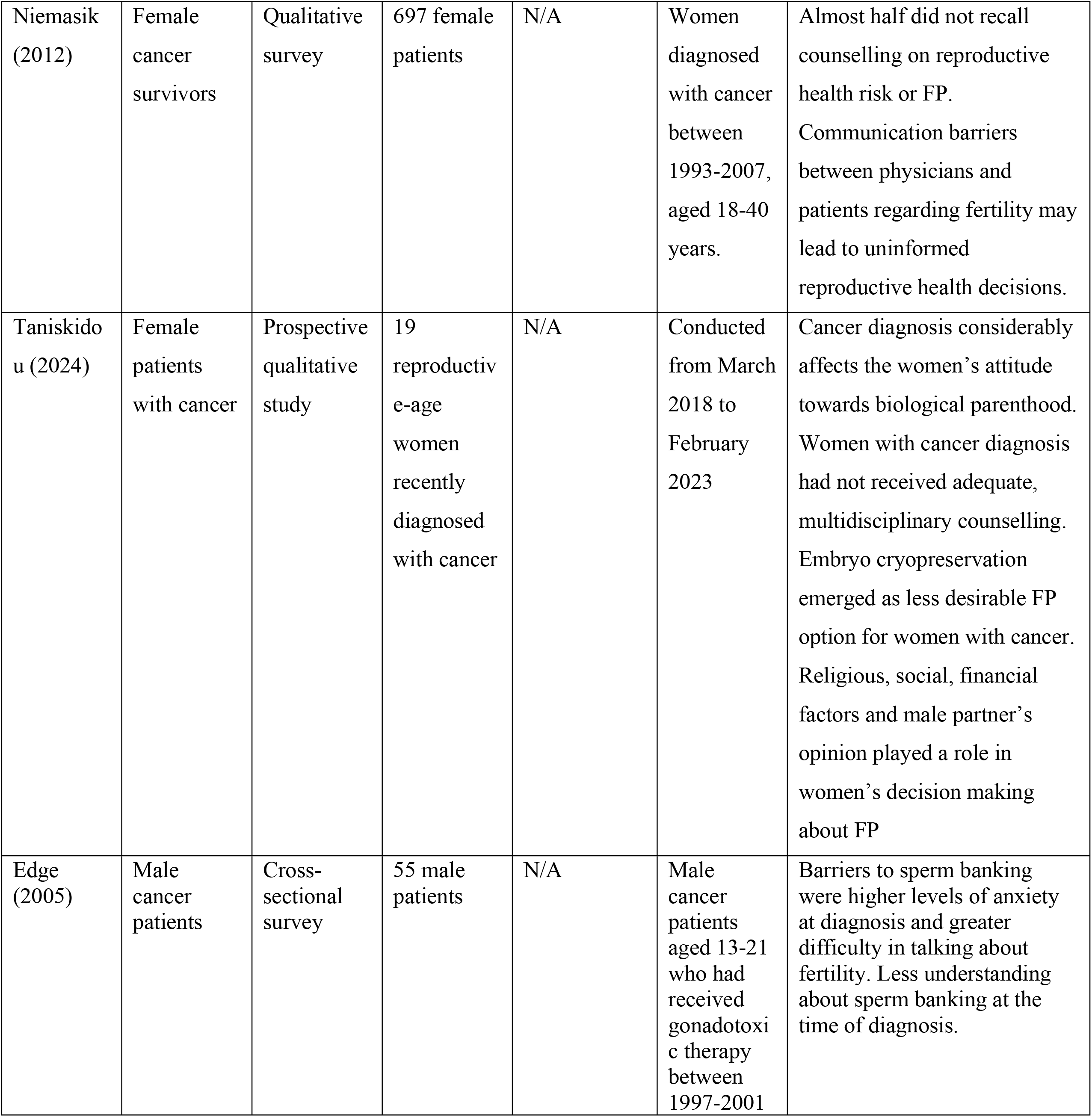
Studies included in this systematic review.

### Knowledge of Fertility Preservation

Awareness of fertility preservation options was consistently low among cancer patients. Studies revealed that between 40% to 70% of participants reported not receiving adequate information on fertility risks before starting treatment [27, 28]. A U.S.-based survey by Quinn et al. found that fewer than half of young adult cancer patients recalled having a fertility discussion with their oncologist [27]. Similarly, Armuand et al. reported that women were significantly less informed than men regarding fertility-related risks, highlighting a gender disparity in information dissemination [16].

### Attitudes and Preferences

Despite limited knowledge, the majority of patients expressed strong interest in preserving fertility, particularly among younger individuals, those without children, and those in long-term relationships [29, 30]. For instance, Peate et al. documented that nearly 80% of women with breast cancer expressed regret about not receiving fertility-related information [31]. Patients viewed fertility preservation as essential to maintaining their identity, life goals, and hope for the future [32]. Cultural and personal values also played a role; in studies from Iran and other low-to middle-income countries, fertility was closely tied to marital prospects and social status [33, 34, 35]. However, in sub-Saharan Africa, a study from Uganda explored shared decision-making about future fertility among parents of children diagnosed with cancer. It highlights how fertility preservation is under-recognized, and parental desires for preserving fertility are deeply entwined with psychosocial considerations in a limited-resource setting [36].

### Barriers to Fertility Preservation

Barriers were both structural and psychosocial. Common challenges included a lack of provider-initiated discussions, high costs of preservation procedures, urgency to begin cancer treatment, and a lack of nearby fertility clinics [15, 35, 37]. Psychological distress at diagnosis often made it difficult for patients to process fertility information [15, 29]. Studies from Canada and the UK emphasized logistical constraints such as unclear referral pathways and insufficient coordination between oncology and fertility teams [38, 39]. Furthermore, cultural taboos and stigma— especially in conservative societies—discouraged open discussions about reproductive health [40, 41].

## Discussion

This systematic review underscores significant gaps in knowledge, counselling, and access to fertility preservation services among cancer patients receiving chemoradiotherapy. This observation is similar to what was reported by Black BP et al. in a thematic synthesis of 29 qualitative studies that found consistent deficits in fertility preservation (FP) knowledge and provider-initiated discussions, compounded by barriers in counselling and decision support [42]. Similarly, Jones G and colleagues found that information gaps, inadequate counselling, limited referrals, and unequal access to fertility preservation, particularly among patients undergoing chemo-radiotherapy [42]. Although the technological capacity for fertility preservation has advanced considerably, patient awareness and utilization remain low due to multifaceted barriers, especially in low-and middle-income countries [36].

The findings affirm previous reviews suggesting that provider communication is pivotal in facilitating access to oncofertility services. Clinicians often face time constraints, limited training, or discomfort when discussing fertility, especially in the context of a life-threatening illness. As a result, patients—especially younger and female—are frequently left uninformed. This lack of counselling violates professional guidelines issued by ASCO and ESMO [1, 2], which mandate fertility discussions at diagnosis.

Cost remains a major deterrent globally. Even in high-income settings, fertility preservation procedures such as oocyte or sperm cryopreservation are often not covered by insurance. The reports from two studies in high-income countries showed that most patients must pay USD 10,000–13,000 (oocyte) or USD 13,000–16,000 (embryo), while coverage for iatrogenic infertility is generally not mandated—only a minority of U.S. states require fertility preservation coverage after cancer treatment [44, 45]. In low- and middle-income countries, the financial burden is even more pronounced, making fertility care inaccessible to the majority. In their study, Black BP and colleagues concluded that high out-of-pocket costs and the absence of insurance reimbursement significantly limit uptake of fertility preservation even in developed healthcare systems [42]. Policy reforms and public funding are needed to bridge this equity gap.

Cultural beliefs further complicate access. In conservative societies, reproductive discussions may be taboo, particularly among unmarried women. In a recent study done by Ghorbani B and colleagues in Iranian female cancer patients, fertility and reproductive discussions were considered private or taboo, especially when done outside marriage [40]. This stigma discourages patients from seeking information or prioritizing fertility. Thus, culturally sensitive counselling is essential. Additionally, mental health support at diagnosis can help patients cope and engage more meaningfully in fertility-related decision-making.

The review also highlights a lack of standardized referral pathways and institutional support for fertility preservation. Integrating fertility services within cancer care pathways and establishing oncofertility coordinators can streamline processes. Educational interventions targeting both healthcare providers and patients have demonstrated improved knowledge, attitudes, and uptake.

Finally, the role of digital health tools, such as online decision aids and telehealth counselling, is increasingly recognized in overcoming geographic and logistical barriers. These tools, if culturally adapted, can empower patients and enhance shared decision-making.

## Conclusion

Despite the recognition of fertility preservation as a survivorship priority, significant barriers persist in patient education, clinician practice, financial access, and systemic coordination. Addressing these challenges requires a multidimensional approach involving policy advocacy, clinical training, infrastructural investment, and culturally competent care. Future research should focus on implementation strategies and outcome evaluation of integrated oncofertility programs.

## Data Availability

All relevant data are within the manuscript and its Supporting Information files

## Acknowledgments

We acknowledge all gynecologic Oncologists working at Kenyatta University, Moi Teaching and Referral Hospital, and Kenyatta National Hospital.

## Ethical Approval

Not applicable for this review.

## Strengths

This systematic review has several strengths. It is the first to synthesize evidence from a wide range of geographic and cultural contexts specifically focused on the knowledge, attitudes, and barriers to oncofertility services among cancer patients undergoing chemoradiotherapy. The use of a rigorous search strategy across multiple databases, a dual-reviewer screening and extraction process, and critical appraisal using the MMAT tool all contribute to the validity of findings. Moreover, the inclusion of both qualitative and quantitative studies allowed for a comprehensive and nuanced understanding of patient experiences.

## Limitations

However, the review has limitations. Only studies published in English were included, which may introduce language bias and limit generalizability. Additionally, grey literature and conference proceedings were not systematically searched, possibly excluding relevant data. The heterogeneity in study designs and outcome measures prevented meta-analysis, reducing the ability to quantify effects. Lastly, although attempts were made to include recent and diverse evidence, the number of included studies remains relatively small, particularly from low- and middle-income countries, where barriers to fertility services may be more pronounced.

